# A continuous indicator of food environment nutritional quality

**DOI:** 10.1101/2021.11.24.21266841

**Authors:** Iris C. Liu, Kayla de la Haye, Andrés Abeliuk, Abigail L. Horn

## Abstract

Food environments can profoundly impact diet and related diseases. Effective, robust measures of food environment nutritional quality are required by researchers and policymakers investigating their effects on individual dietary behavior and designing targeted public health interventions. The most commonly used indicators of food environment nutritional quality are limited to measuring the binary presence or absence of entire categories of food outlet type, such as ‘fast-food’ outlets, which can range from burger joints to salad chains. This work introduces a summarizing indicator of restaurant nutritional quality that exists along a continuum, and which can be applied at scale to make distinctions between diverse restaurants within and across categories of food outlets. Verified nutrient data for a set of over 500 chain restaurants is used as ground-truth data to validate the approach. We illustrate the use of the validated indicator to characterize food environments at the scale of an entire jurisdiction, demonstrating how making distinctions between different shades of nutritiousness can help to uncover hidden patterns of disparities in access to high nutritional quality food.

**CCS CONCEPTS:**

- **Applied computing → Life and medical sciences**.

## 1 INTRODUCTION

Initiatives that improve diets are needed [22, 30, 37] as poor diets led to an estimated 11 million deaths globally in 2017 [13], and are a key cause of major chronic diseases including obesity, cancer, and heart disease [42]. Emerging evidence has shown that foodv environments—the physical and social spaces in which people acquire and consume food—can profoundly impact diet and related diseases [16, 37]. The key mechanisms proposed to explain the relationship between food environments and unhealthy eating behaviors are that (i) low access to healthy food options can induce unhealthy choices out of convenience or necessity [10, 11, 40], and (ii) exposure to an abundance of unhealthy options can cue or trigger unhealthy eating [14, 39, 41].

Effective, robust measures of food environment quality are required by researchers and policymakers investigating the effects of the food environment on individual dietary behavior and designing targeted public health interventions. For both applications, there is great value in a single indicator that can provide a summary assessment of nutritional quality in a single score that can be applied to characterize diverse neighborhoods and food environments, at scale [36].

The most commonly used indicators of food environment nutritional quality, *food deserts* [40] and *food swamps* [14], are limited in scope, measuring the binary presence or absence of single categories of food outlet type. Neighborhoods are characterized as food deserts if there are few or no healthy food outlets in a range of proximity (e.g., 0.5 or 1 mile) of residents living in a neighborhood, where healthy outlets are operationalized as supermarkets [27]. Food swamps are defined as neighborhoods that have a high percentage of unhealthy food outlets relative to all total outlets, or relative to healthy food outlets, where unhealthy food outlets are operationalized as fast-food or fast-food and convenience stores [14, 25]. While food sold at fast-food outlets, supermarkets, and convenience stores has been demonstrated to have a meaningful relationship on nutritional health [12, 15, 23], the focus on a single type of outlet is limited given the great variability in types of food outlets and the nutritional quality menu offerings within each type.

Fast-food restaurants are a prime example of the continuum of nutritional quality of menu offerings. Restaurants are commonly identified as fast-food if they fall under a certain category in the North American Industry Classification System (NAICS) [28], that of limited-service restaurant [14, 40] (code 722513). This classification system was developed over 20 years ago when most limited-service restaurants served a similar type of American menu, e.g. burgers, fries, and shakes. There has been a shift in the food offered at limited-service restaurants over the past couple of decades, including the introduction of fast and healthy foods such as salads; ‘bowls’; and non-fried beans, rice, and vegetable dishes. However, measures and classification systems have not yet caught up, and restaurants falling under NAICS code ‘limited-service restaurant’ range from McDonald’s to the salad chain Sweetgreen. A robust, *continuous* indicator of food environment nutritional quality is needed, that moves beyond the binary focus on the prevalence of outlets within single categories of food outlets, to capture the gradation of the nutritional quality within and across categories.

### 1.1 Related work

While an indicator-based approach has not been previously developed at the level of a food outlet, neighborhood, or food environment, quantified and continuous indicators have been designed for evaluating the nutritional quality of an individual food item or diet based on its nutritional components. These indicators focus on the quality and balance of diverse nutritional components rather than single out the total amount of energy or calories, since it is well established that the caloric content of a food is not correlated with recommended nutrient availability, and that poor nutrition is caused not only by consuming high-calorie foods, but from over-consuming low-nutrient foods [20, 26, 38].

Nutritional quality indicators combine multiple nutrient or dietary components simultaneously, on a density basis. One type of approach, the Healthy Eating Index (HEI), relies on the amounts of key dietary components (i.e., total fruit, whole fruit, legumes, etc.) [21]. Another type of approach, nutrient profile density scoring, requires the amounts of individual macro and micronutrients (i.e., protein, sodium, calcium) composing a food item. Both types of approaches have been demonstrated to be reliable and valid tools for quantifying nutritional quality [17, 24, 35].

A couple of analyses have applied the HEI to summarize the quality of the food environment at a limited scale. Reedy et al. (2015) suggest applying the HEI to the retail food environment, illustrating the approach on a sample dollar menu from a fast-food restaurant [32]. Hearst et al. (2013) applied the HEI to 8 fast-food outlets in a study analyzing trends in nutritional quality over time [18]. Meanwhile, nutrient density scoring of individual food items has been used in multiple regulatory applications, including evaluating labeling and marketing of snack foods, food tax programs, and defining school food standards [1, 9, 19, 24, 29, 34]. A recent study used nutrient density scoring across a larger sample of food items sold at 700 chain restaurants to illustrate the absence of a linear relationship between caloric and nutrient content in food items [20].

However, nutrient density scores have yet to be deployed to characterize restaurants across food environments, at scale. The absence of an approach developed at scale may be due to measurement, in that it has been difficult to assess the nutrients available in the millions of menu offerings across the hundreds of thousands of food outlets across the food environment. In this paper, we show that this is now possible, given the wide availability of digital menu data, and the introduction of an approach to estimate nutrients for given menu items.

### 1.2 Overview

This work contributes to the ability to characterize the nutritional quality of diverse food environments across a continuum and at scale. We focus on the retail restaurant food environment, where the majority of Americans’ food budget is spent [33]. We introduce a summarizing indicator of *restaurant nutritional quality*, RNQ, that takes in descriptions of food items from a restaurant’s menu, estimates their nutrient values, and returns an overall score of the restaurant’s nutritional density across all menu items. The RNQ measure can be applied to compare restaurants within a food outlet category, or across categories. Verified nutrient data for a set of 522 chain restaurants from the nutrition data company Nutritionix is used as ground-truth data to validate the approach.

We illustrate the use of the RNQ measure to characterize food environments at the scale of a jurisdiction, focusing on restaurants falling within the same food outlet category. The example demonstrates how making distinctions between different shades of nutritiousness within a category can help to uncover hidden patterns of disparities in access to high nutritional quality food.

The paper is organized as follows. We develop the restaurant-level RNQ indicator, and describe the validation analysis using ground-truth nutrient information, in Methods. The Results section presents findings of the validation analysis and an example illustrating the use of the continuous indicator of restaurant nutritional quality in the food environment of the greater Los Angeles area. In the Discussion, we interpret findings and discuss future work, which will scale the approach introduced here across food outlet categories and environments.

## 2 METHODS

### 2.1 Approach

We approach the development of a restaurant-level indicator across three key steps: (1) obtaining a target menu dataset representing restaurant names and descriptions of each menu item, (2) obtaining or estimating the nutritional content of menu items at scale across the entire restaurant food environment, and (3) scoring the nutritional quality of a restaurant.

Step (1), establishing a target restaurant menu dataset to score, may be done by accessing menu data from various software companies that maintain extensive databases of metadata on points of interest, including food outlets and their menus, such as Yelp [8] and Foursquare [5]. Limited selections of restaurant menu items coming only from chain restaurants are available from personal nutrition-logging applications such as Nutritionix [7], Chronometer [4], and MyFitnessPal [6]. In this work, we use restaurant menu data from Nutritionix as our target dataset, as it represents the largest database of verified restaurant menu items and their nutrients, and can thus be used for validation analysis of the proposed method.

Step (2) involves estimating the nutritional content of menu items, i.e., the levels of the macro and micronutrients based on all the composing food ingredients, given the menus and menu item-level descriptions obtained from Step (1). This is necessary to estimate because nutritional content is not provided by restaurants, with the notable exception of chain restaurants with more than 20 outlets, as mandated by the FDA under the Affordable Care Act as of May 2018 [20, 31]. Here, we introduce an approach to characterize the food environment in the absence of nutrient information posted on restaurant menus, enabling quantification of the nutrition of the food environment including and beyond large chain restaurants. We make use of the United States Department of Agriculture (USDA) National Nutrient Database for Standard Reference [2].

This nutrition dataset, curated by the USDA, is the major source of food composition data in the US used by researchers, policymakers, health professionals, and others, and is the starting point for dieticians in scoring the nutritional quality of an individual’s diet [32]. We retrieve matches to food items in the target dataset from this database using its API and built-in language-based matching algorithm. We add additional natural language postprocessing steps to improve the matching based on food item name descriptions.

Step (3) involves developing an aggregate indicator of the nutritional quality of a restaurant based on its menu offerings, the RNQ. We make use of an existing nutritional quality index – nutrient density scoring [34, 35]. We apply nutrient density scoring at the menu-item level, and then aggregate up to the level of an entire restaurant through a median-based statistic.

### 2.2 Restaurant-level indicator of nutritional quality

Here we describe our process for designing an index of the healthfulness of a restaurant based on its menu offerings. This step begins after a target dataset of restaurants and their menu item descriptions as they appear on the menu has been obtained (Step 1).

#### 2.2.1 Preprocessing menu data

There are two main steps in the preprocessing process: data cleaning and data filtering. Data cleaning is applied to remove unnecessary fields while preserving information of interest including restaurant name, restaurant category, menu item name, ingredients, and food category. Menu item strings are also cleaned in this step. To select menu items for the accurate nutritional scoring of single-serving adult meals we apply a set of keyword filters. We remove children’s menus, dishes for sharing, beverages, and single-ingredient foods. This is done to create an index that is comparable across restaurants based on their diversity of offerings.

#### 2.2.2 Estimating the nutritional quality of menu items

After menu item filtering, we fetch food nutritional details of each menu item from the United States Department of Agriculture (USDA) National Nutrient Database for Standard Reference through its queryable API, FoodData Central. It contains nutrient profile information for over 8,000 branded and unbranded food items, each with up to 70 nutrients, although not all are available for each menu item. We access seven nutrient values for calories, protein, carbohydrates, total fat, saturated fat, fiber, and sugar.

For each USDA-matched menu item, there exists a “score” field, which represents a relative score indicating how well the food matches the search criteria. We use the score as a matching threshold. We retain all matched menu items from the USDA database with a score above 100. Sensitivity analysis determined that a matching score threshold of 100 results in the most accurate results, when compared with ground truth data.

Menu items obtained from the USDA database are post-processed to minimize inaccuracies and inconsistencies. We remove menu items with 0 calories, and with missing nutritional values. We rebase extreme outliers, applying a maximum cap limit across nutrient values. We also apply the same set of keyword filters mentioned in Section 2.2.1 to the USDA matched menu items to ensure the matched sample contains only single-serving adult meals.

Finally, to obtain an overall nutrient value content for each nutrient category for the target dataset menu item, we average over the nutrient values of each retained menu item match from the USDA database.

### 2.3 Restaurant nutritional density scoring using menu item nutrients

Nutrient density scoring aggregates the nutritional information of a food or menu item into a single score by measuring the relative balance of recommended to restricted nutrients present. It can be seen as a representation of the overall nutritional quality of a food item.

There are a handful of available nutrient density scoring systems from the nutrition literature, most applying some relationship between the value of recommended nutrients (e.g., protein, fiber) to restricted nutrients (i.e. saturated fat, added sugars, sodium). These approaches range in the number of nutrient constituents they require, whether both recommended and restricted (by USDA guidelines) nutrients are included, and whether a binary or continuous score is produced. A review of such methods is provided in Santos et al. (2021) [32] and Ho et al. (2020) [20]. Of these, two available scoring systems meet the criteria for this study of being (1) based on nutrients available from almost all foods provided in the USDA database and (2) defined along a continuous scale: Scheidt and Daniel’s “Ratio of Recommended to Restricted Nutrients” (RRR) [35] and Fulgoni, Keast and Drewnowski’s “Nutrient Rich Foods” (NRF) score [17].

Of these two methods, we choose to develop our restaurant-level nutrition indicator using the RRR. While both methods are normalized to daily recommended values (DRV) of each nutrient, only RRR balances the levels of recommended and restricted nutrients by setting them in a weighted ratio centered around 1, providing a more interpretable index.

The RRR score is a ratio of percent DRV of six recommended nutrients over five nutrients to restrict per serving, with weights added to balance the contribution of the recommended nutrients and the restricted nutrients. The DRV refers to the U.S. Dietary Guidelines and daily reference values, e.g. daily recommended intake of protein is 50 grams [3].

The original RRR equation contains the recommended nutrients protein, fiber, vitamin A, vitamin C, iron, and calcium; and the restricted nutrients sugars, cholesterol, saturated fat, and sodium [35]. We modify the equation based on the nutrients available in the majority of the USDA data and the majority of menu items in the Nutritionix ground truth data (Section 2.4), 60% of which does not contain values for the 4 micronutrients vitamin A, vitamin C, iron, and calcium. The modified RRR score (RRR-m) is expressed as:

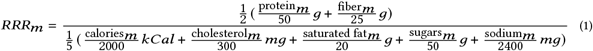

Through the use of the weights and the ratio, the score is balanced such that scores close to 0 represent lower nutrient density; scores equal to 1 indicate an equal proportion of recommended to restricted nutrients; and scores above 1 include more recommended nutrients than restricted nutrients.

#### 2.3.1 Restaurant nutritional quality (RNQ) score

A restaurant’s nutritional quality (RNQ) score is computed as the median of the RRR-m scores across all menu items within that restaurant, as sensitivity testing determined this statistic to produce more accurate RRR-m scores, compared with ground truth data (Section 2.4) than the mean. To ensure sufficient items for stable estimates of restaurant scores, we do not attempt to obtain a restaurant-level score for restaurants with less than 5 menu items, after all filters have been applied. Sensitivity analyses determined restaurant-level RRR-m scores were robustly accurate for restaurants with ≥ 5 menu items.

### 2.4 Validation Analysis

#### 2.4.1 Data and preprocessing

We collect a dataset providing the nutrient profiles for menu items, Nutritionix, as ground-truth for validation and calibration analysis. Nutritionix is a software company that aggregates and manages nutrition data on menu items from a large selection of chain restaurant brands, retail foods, and common foods for use by personalized health mobile applications and food industry websites. The database consists of 38,275 menu items across 1,436 restaurant chain brands in the United States, organized at the level of menu items sold by each chain. For each menu item within a brand, we obtain information on its name (as a phrase), calories, and values of the macronutrients sodium, protein, dietary fiber, saturated fat, cholesterol, and sugars.

To create a dataset fit for RNQ scoring, we implement the data post-processing steps applied to matches from the USDA database described in Section 2.2.2, using keywords to filter out items that do not represent single-serving adult meals, removing items with 0 calories or missing nutrient values, and rebasing extreme outliers. We retain for scoring only restaurants with ≥5 menu items, losing 35.5% of restaurants due to this filter. After all reductions, 7,013 menu items from over 522 restaurant brands remain in the dataset for ground truth analysis.

#### 2.4.2 Comparative analysis with ground-truth data

Our validation analysis focuses on comparing RNQ scores for each of the 522 restaurant brands in the post-processed Nutritionix database obtained using estimated nutrient values and the ground truth nutrient values for each of the 7,013 menu items. We estimate the nutrient values for each target menu item using the approach in Section 2.2.2, fetching menu-item matches from the USDA database, and post-processing the matched menu item data. We then obtain an RRR-m score for each menu item and an RNQ score for each restaurant brand, which we call the the RNQ-USDA, as described in Section 2.3. Separately, using the nutrient values provided in the ground truth dataset, we obtain an RRR-m score for each menu item and an RNQ score for each restaurant brand, which we call the RNQ-GT. We model the linear correlation between the RRR-m score and the RRR-GT score at the menu-item and restaurant brand level.

## 3 RESULTS

### 3.1 Ground truth analysis

We present the results of the ground truth analysis on the Nutritionix dataset, first at the menu-item level, and then at the restaurant brand level for which the measure is designed to be used.

#### 3.1.1 Menu-item level nutritional scoring

Table 1 illustrates effective and ineffective menu item matches between the Nutritionix ground truth dataset and the USDA database. It presents the three menu items with the lowest error and the highest error between the RRR-m score calculated on the estimated nutrient values and ground truth nutrient values. Error is calculated as the ratio of the difference between the estimated and ground-truth RRR-m over the ground truth RRR-m. Low error menu item scores are achieved with matches to exact phrases, such as the exact match on the menu item ‘peanut noodles’, or matches between sets of items with similar nutrient profiles, like the match between the Nutritionix menu item ‘Mrs. Fields Cookies, Semi-Sweet Chocolate’ and various Mrs. Fields cookie products that have a similar composition of sugar and saturated fat. Low error menu item RRR scores may also be achieved by averaging the nutrient values over multiple menu items matching different substrings. For example, the target item ‘Double Smoked Cheddar Cheese Slider’ from the Nutritionix database does not find an exact match in the USDA database, but find matches to items in the USDA database including the substrings ‘Double Smoked’, ‘Cheddar Cheese’, and ‘Cheddar Slider’; averaging across these items converges on an accurate RRR-m score.

**Table 1:** Effective and ineffective name matches between the Nutritionix dataset and the USDA database.

| Original item name | Low Error |  |  |  | Original item name | High Error |  |  |
| --- | --- | --- | --- | --- | --- | --- | --- | --- |
|  | Matched item names | RRR score, ground truth | RRR score, estimated |  |  | Matched item names | RRR score, ground truth | RRR score, estimated |
| Mrs. Fields Cookies, Semi-Sweet Chocolate | ['MRS. FIELDS, COOKIE',<br>'MRS. FIELDS, CHOCOLATE CHIP COOKIES',<br>'MRS. FIELDS FROZEN VANILLA DAIRY DESSERT BETWEEN TWO MRS. FIELDS CHOCOLATE CHIP COOKIES SANDWICHES, MRS. FIELDS',<br>'MRS. FIELDS, BUTTER PECAN PRALINE COOKIES',<br>'MRS. FIELDS, COOKIES, OATMEAL RAISIN WITH WALNUTS',<br>'MRS. FIELDS, CHOCOLATE CUPCAKES',<br>'MRS. FIELDS, COOKIES, SEMI-SWEET CHOCOLATE CHIP'] | 0.23 | 0.23 |  | French Lentil & Kale Large | ['FRENCH LENTILS',<br>'FRENCH LENTILS ORGANIC SOUP',<br>'KALE & LENTIL SOUP',<br>'LENTIL, POTATO & KALE ORGANIC SOUP, LENTIL, POTATO & KALE',<br>'FRENCH LENTIL SOUP',<br>'ZIYAD, LARGE WHOLE LENTILS'] | 1.36 | 12.55 |
| Double Smoked Cheddar Cheese Slider | ['DOUBLE SMOKED CHEDDAR CHEESE',<br>'TURKEY & SMOKED CHEDDAR SLIDERS',<br>'SMOKED TURKEY AND DOUBLE CHEESE FLATBREAD',<br>'HONEY TURKEY & CHEDDAR CHEESE SLIDERS',<br>'DOUBLE SMOKED SAUSAGES',<br>'HAM & CHEDDAR SLIDERS',<br>'TURKEY & CHEDDAR SLIDERS'] | 0.77 | 0.77 |  | Crisp Pinto Bean Burrito | ['NATURAL PINTO BEAN & CHEESE BURRITO',<br>'PINTO BEANS'] | 1.14 | 10.76 |
| Peanut Noodles | ['PEANUT NOODLES'] | 0.57 | 0.57 |  | Spanish Rice | ['SPANISH RICE',<br>'SPANISH RICE PILAF MIX',<br>'SPANISH RICE'] | 10.67 | 0.79 |

Food items such as “French Lentil & Kale Large”, “Crisp Pinto Bean Burrito”, “Spanish Rice” show the highest errors using our approach. We observe that good matches on language are not sufficient for determining a good match to the actual food item. A food item matched on a substring may have fewer ingredients than the target food, as in the target item ‘Crisp Pinto Bean Burrito’ matched to the single ingredient ‘Pinto Beans’ in the USDA database. The matched item can also have more ingredients, as in the target item “French Lentil & Kale Large” matched to a French lentil and kale soup dish, and a French lentil, kale, and potato dish in the USDA database. In addition, because menu item descriptions do not comprehensively list out all ingredients, a menu item with the same name may be prepared with multiple variations of ingredients at different restaurants, such as ‘burger’.

Fig 1 shows the correlation between the menu item level RRR-m scores calculated using ground truth nutrient values and the estimated nutrient values. Our approach demonstrates an approximately 50% (Pearson R=0.51) agreement with validation data in identifying menu items.

**Figure 1:**
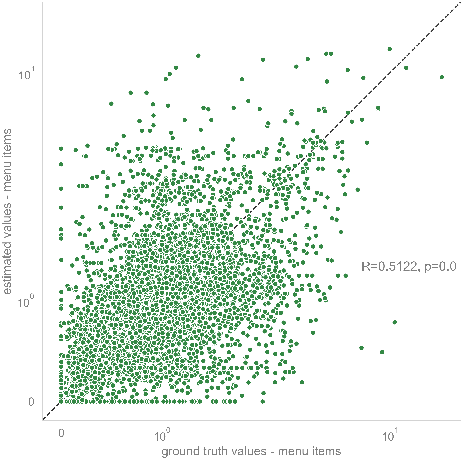
Menu-item level correlation between ground truth and estimated nutritional density (RRR-m) scores.

#### 3.1.2 Restaurant brand-level nutritional scoring

Fig 2 shows the correlation between restaurant brand-level RNQ scores utilizing the ground truth vs. the estimated nutrient values. The estimated and ground-truth RNQ scores are positively correlated with a Pearson correlation coefficient of 0.83, with statistical significance. This result suggests that while our method may not be sufficiently accurate at scoring the nutrition of individual menu items, by taking the median over all menu items in a restaurant brand, we can achieve a reasonably accurate restaurant brand-level score.

**Figure 2:**
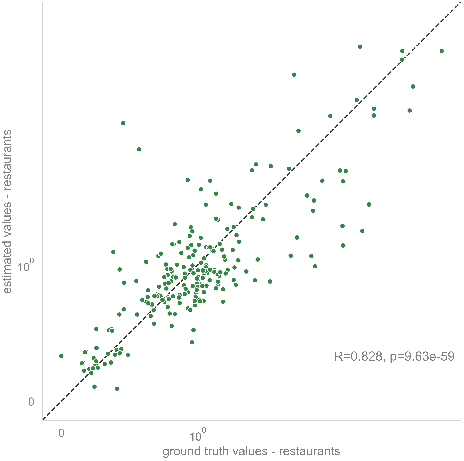
Restaurant brand-level correlation between ground truth and estimated restaurant nutritional quality (RNQ) scores.

Fig 3 shows histograms of the distribution of the RNQ scores across the 522 Nutritionix restaurant brands calculated on the ground truth and estimated nutrient values. To quantify the similarity between the distributions we measure the Kullback-Liebler Divergence (KLD), and compare it with the KLD between the ground truth distribution and the uniform distribution. The KLD between the ground truth and estimated values is 0.01, whereas the KLD between the ground truth distribution and the uniform distribution is 0.12, more than 10 times larger, suggesting the distributions are closely matched. A vertical line at *RNQ* = 1 in Fig 3 indicates whether restaurant brands serve food with a greater balance of recommended nutrients to restricted nutrients (*RNQ >* 1) or a greater balance of restricted to recommended nutrients (*RNQ <* 1). The estimated percentage of restaurant brands with a higher balance of recommended to restricted nutrients is 37%, compared with 43% when using the ground-truth nutrient values.

**Figure 3:**
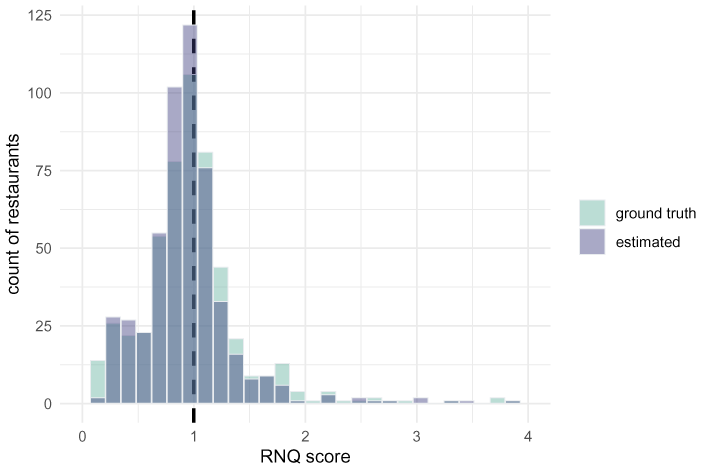
Overlapping histograms of the distribution of estimated and ground truth RNQ scores across the 522 restaurant brands.

Table 2 shows restaurant brands and their menu items for the top three highest, bottom three lowest, and median three restaurant brands based on their estimated RNQ scores. We observe that restaurant brands with the highest RNQ scores feature menu items that are not necessarily low in calories, but have a high proportion of the recommended nutrients protein (from animal and plant sources), and fiber (from whole grains and vegetables). The lowest RNQ scores come from restaurants serving primarily “low-nutrient energy-dense” items that are high in added sugars and saturated fat, mostly dessert items. Middle-RNQ scores come from restaurant brands serving more diversified menus including a mix of healthier items items high in protein and/or fiber (wraps, salads, and bowls), items higher in saturated fat (wrap with mayonnaise-based dressings or cheese), and items high in carbohydrates but low in protein or fiber (muffins, white rice).

**Table 2:** Top 3 restaurants with the highest, middle, and lowest estimated RNQ score according to estimated nutrient values. The table shows the RNQ score based on estimated nutrient values (RNQ-USDA) and ground truth nutrient values (RNQ-GT) for a sample of 5 menu items per restaurant.

| Highest RNQ |  |  | Middle RNQ |  |  | Lowest RNQ |  |  |
| --- | --- | --- | --- | --- | --- | --- | --- | --- |
| Restaurant | RNQ-USDA | RNQ-GT | Restaurant | RNQ-USDA | RNQ-GT | Restaurant | RNQ-USDA | RNQ-GT |
| **GRK Fresh Greek** | 3.51 | 1.56 | **Just Fresh** | 0.82 | 0.99 | **Whitey's Ice Cream Grocery** | 0.23 | 0.24 |
| Tzatziki Sampler with 2 whole wheat pitas | 3.28 | 1.12 | Cranberry Orange Muffin | 0.45 | 0.35 | Mint Chocolate Chip Ice Cream | 0.23 | 0.24 |
| Plate - Chicken with whole wheat pita | 3.51 | 1.82 | Western Omelet | 0.50 | 1.09 | Mississippi Mud | 0.30 | 0.18 |
| Chicken Soup with whole wheat pita | 3.46 | 1.85 | Small Caesar Salad | 0.82 | 0.55 | Ice Cream, Swiss Almond Chocolate | 0.21 | 0.24 |
| Pita Yeero - Lamb & Beef whole wheat pita | 3.51 | 1.86 | Huevos Wrap | 0.91 | 0.99 | Ice Cream, Moose Tracks | 0.21 | 0.22 |
| Pita Yeero - Chicken with whole wheat pita | 3.51 | 1.91 | Santa Fe Chicken | 1.01 | 1.84 | Chocolate Chip Ice Cream | 0.19 | 0.23 |
| **Protein House** | 3.33 | 1.71 | **Crazy Bowls & Wraps** | 1.20 | 1.73 | **Good Times Burgers & Frozen Custard** | 0.26 | 0.25 |
| Chicken Caesar Wrap | 1.22 | 1.70 | Caesar Wrap | 1.22 | 1.59 | Big Daddy Bacon Cheeseburger | 0.86 | 0.98 |
| Southwest Veggie Wrap | 1.44 | 1.58 | Crispy Chicken Bites | 1.25 | 1.66 | Breakfast Burrito | 0.87 | 0.63 |
| Portobello Sandwich with Toasted Ezekiel Bread | 3.33 | 3.60 | Pesto Bowl | 1.08 | 1.80 | Chocolate Peanut Butter Crunch Spoonbender, Medium | 1.36 | 0.25 |
| Power Wrap | 4.32 | 1.71 | Lobster Rangoon | 0.71 | 0.45 | Onion Rings | 0.58 | 0.57 |
| Thai Monster Bowl with Whole Wheat Pasta | 5.30 | 3.10 | Thai Wrap | 1.08 | 1.80 | Mac & Cheese with Green Chile | 0.70 | 0.83 |
| **Veggie Patch** | 2.62 | 2.47 | **District Taco** | 1.04 | 1.44 | **Dunkin' Donuts** | 0.29 | 0.41 |
| Garden Broccoli Bites | 2.31 | 1.66 | Garlic-Lime Rice | 0.66 | 0.64 | Chocolate Chip Muffin | 0.28 | 0.28 |
| Crispy Chik'n Cutlet, Meatless | 2.61 | 2.63 | Huevos Rancheros | 0.88 | 1.17 | Maple Creme Donut | 0.47 | 0.34 |
| Falafel Chickpea Balls | 2.63 | 2.47 | Quesadilla - Chicken | 1.22 | 1.50 | English Muffin | 1.85 | 1.50 |
| Chik'n Bites, Buffalo | 2.76 | 2.45 | Burrito Mojado - Chicken | 1.67 | 1.38 | Vanilla Creme Donut | 0.16 | 0.32 |
| Classic Falafel, Chickpeas & Spices | 2.96 | 2.47 | Vegetarian Black Beans & Rice | 2.72 | 2.80 | Apple Crumb Donut | 0.25 | 0.37 |

### 3.2 Illustrative application

We now illustrate the application of the nutritional density scoring system to characterize the variability in nutritional quality of restaurant brands across a food environment. Using a database of 40,000 restaurant outlets in the greater Los Angeles area (LA) and their spatial locations obtained from Foursquare via their public API, we match the scored Nutritionix restaurant chain brands by name to find their locations in LA. Of the 522 chain brands in the Nutritionix database, 174 brands are found in the Foursquare database, with 6,078 physical outlets across LA. All of these restaurant chain brands fall within the same NAICS category of ‘limited-service restaurant’ (code 722513) [28]. In the following, we illustrate the nutritional diversity across restaurants grouped within this category. The subsample of 174 restaurant chain brands have RNQ scores ranging from 0.15 to 2.6. We focus on the highest and lowest-scoring brands according to their ground truth nutrient values. Specifically, we designate the “most nutritious” brands as those with RNQ scores greater than 1.3, representing the top 7% of the sample of 174 restaurant chains, and the “least nutritious” as those with RNQ scores < 0.3, representing the bottom 7% of the restaurant chain sample. Notably, the top 7% of restaurant brands by RNQ score represent only 92, or 1.5%, of the sample of 6,078 physical outlets. Examples include Sweetgreen, a salad chain; Veggie Grill, which serves plant-based salads, bowls, and sandwiches; and Rubio’s Coastal grill, which features a seafood and vegetable-focused taco and burrito menu. The bottom 7% of restaurant brands by RNQ score represent 357 food outlets across the LA food environment, 6% of the sample. Examples include McDonald’s, Winchell’s doughnut house, and Baskin-Robbins.

The map in Fig 4 shows the locations of these least and most nutritious food outlets in the LA area, demonstrating an unbalanced distribution. The least nutritious outlets are found everywhere, apparently distributed uniformly at random. In stark contrast, the most nutritious outlets are not uniformly distributed but are clustered in specific neighborhoods; in particular, the affluent west to east corridor in the north of the LA area between Santa Monica, Beverly Hills, and downtown Los Angeles.

**Figure 4:**
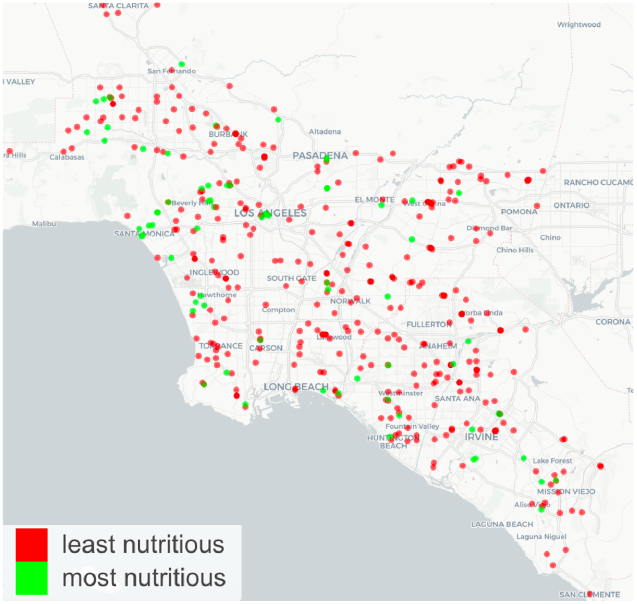
Map of the greater Los Angeles area indicating locations of most nutritious (green, *RNQ >* 1.3) and least nutritious (red, *RNQ <* 0.3) restaurant outlets scored from the Nutritionix dataset.

## 4 DISCUSSION

In public health nutrition research, individuals’ access and exposure to healthy and unhealthy food environments, and the effect on the healthiness of their food choices, is a major area of research that informs public health policy. The most commonly used indicators of food environment nutritional quality are based on the presence or absence of entire categories of food outlets, e.g. *food deserts* and supermarkets, or *food swamps* and fast-food. This work is the first to introduce an index of restaurant nutritional quality that exists along a continuum, that is also deployable at scale across food environments. We use a large ground truth dataset consisting of menus items and corresponding nutrient information for over 500 restaurant chain brands from the personal nutrition software company Nutritionix to validate the approach, obtaining a restaurant nutritional quality (RNQ) score for each restaurant, and comparing the estimated scores to scores calculated using the ground truth nutrient profile information. While the approach does not return reliable matches at the menu-item level, at the restaurant level it is designed for, the approach achieves a reasonably accurate correlation of 0.83 between estimated and ground truth values due to laws of averages.

In an illustrative example, we apply the RNQ scoring measure to the restaurant chain brand restaurants included in the Nutritionix dataset to find their spatial placement across the food environment of the greater Los Angeles area. The example demonstrates the value of moving beyond the use of single-category indices to characterize the healthiness of the food environment, to quantifying the range of nutritional options available. First, a wide distribution of nutritional densities is demonstrated across restaurants categorized under the single category of ‘limited service’ restaurant (NAICS code 722513), a business listing code often used to define ‘fast-food’ outlets. When the RNQ scoring indicator is applied beyond the Nutritionix dataset, the distribution of nutritional density scores will likely become even wider. Second, analysis of the placement of the physical outlets of these restaurant chain brands across the LA food environment demonstrates clear patterns of disparity in access to nutritious food environments. Restaurant outlets on the more nutritious end of the spectrum are closely clustered in specific neighborhoods, while over-saturation of outlets on the least nutritious end of the spectrum appears to be a homogeneous phenomenon.

The measure comes at a time when the need for a continuous indicator of food outlet and environment quality is in high demand, as data capture on individual-level exposure to food environments is becoming increasingly used in both small-sample nutrition studies and population-wide analyses. Access to a fine-grained measure that can quantitatively characterize specific types of food environment, at scale across study populations, can transform the way we measure and characterise exposures and access to specific nutrients. In ongoing work we are using the restaurant-level indicator as the basis for an indicator of the nutritional quality of neighborhood food environments. This requires obtaining a dataset of menus from restaurants across the food environment; we are currently seeking menu data for this purpose from Yelp’s API. Future analyses will seek to compare the continuous indicator of food environment nutritional quality to commonly-used indicators including food desert and food swamp status, in their relationship to socio-demographics and diet-related health outcomes.

## Data Availability

The code produced and data used in this study are available on our GitHub page at https://github.com/irisliucy/ckids-project-usc

## ACKNOWLEDGMENTS

We would like to thank Esteban Moro and Mohsen Bahrami at MIT Media Lab for preparing and sharing the Foursquare restaurant database used in the illustrative example. We would also like to thank Spoorti Nidagundi for her early work on this project. The authors would like to acknowledge the USC Center for Knowledge-Powered Interdisciplinary Data Science (CKIDS) for providing the team infrastructure for developing this project.

